# Effects of Pantoprazole Based on Dialysis Status: A Protocol and Statistical Analysis Plan for a Secondary Analysis of the REVISE Trial

**DOI:** 10.64898/2026.11.02.26367075

**Authors:** Alex Thabane, Oleksa Rewa, Diane Heels-Ansdell, Sean M. Bagshaw, Rachel Jeong, Mark Borthwick, Neill KJ Adhikari, Fernando G. Zampieri, Marlies Ostermann, John Muscedere, John Marshall, Adam Deane, Francois Lauzier, Yaseen M. Arabi, Akira Kuriyama, Karen Burns, Christopher J. Yarnell, Gloria Vázquez Grande, Nicole Zytaruk, Deborah Cook, the REVISE Investigators, Canadian Critical Care Trials Group, and the Australian and New Zealand Intensive Care Society Clinical Trials Group

## Abstract

**Background:** A recent international randomized trial found that the proton pump inhibitor (PPI) pantoprazole significantly reduced the risk of clinically important upper gastrointestinal (GI) bleeding among invasively ventilated patients in the intensive care unit (ICU), with no effect on mortality. Pantoprazole may be of particularly high utility among patients at high-risk of bleeding, such as those receiving renal replacement therapy (RRT) prior to ICU admission. However, given the complex mechanisms by which renal insufficiency may increase bleeding risk, as well as the possible association between PPI use and adverse kidney outcomes, investigating whether kidney failure treated with RRT at baseline modifies the treatment effect of PPIs is clinically relevant.

**Methods:** We have planned a *post hoc* secondary analysis of the Re-evaluating the Inhibition of Stress Erosions (REVISE) Trial (NCT03374800). The primary objective if this study is to evaluate whether any RRT on study day 1 (e.g., maintenance RRT in the outpatient setting, or RRT initiated in the hospital [intermittent hemodialysis, continuous renal replacement therapy, sustained low efficiency dialysis, or peritoneal dialysis]) modifies the treatment effect of pantoprazole on clinically important upper GI bleeding and 90-day mortality. The secondary objective is to evaluate whether maintenance RRT (intermittent hemodialysis or peritoneal dialysis) in an outpatient setting before the index hospital admission modifies the effect of pantoprazole on clinically important upper GI bleeding and 90-day mortality. Secondary outcomes for both analyses are patient important upper GI bleeding, ICU and hospital mortality. We will perform Cox proportional-hazard analyses, including the RRT exposure of interest, randomized group, and their interaction for each outcome, reporting results as hazards ratios with 95% confidence intervals, and p-values for the interaction term.

**Conclusion:** Results of this study will inform critical care practice, particularly for the care of patients at risk of stress-related GI ulceration in the ICU who have received RRT.

## INTRODUCTION

A large population-based study of 531,090 consecutive adult (≥18 years) residents of Ontario, Canada admitted to the intensive care unit (ICU) showed that one in four patients had preexisting chronic kidney disease, which was associated with adverse health outcomes and high healthcare resource consumption [1]. In critically ill patients, the incidence of acute kidney injury (AKI) is estimated to range from 35% to 57% [2–5]; of these patients, approximately 3-24% will have AKI severe enough to receive renal replacement therapy (RRT), with the prevalence particularly high among those receiving invasive ventilation [4, 6] and in low-resource settings [5].

Given that kidney failure and exposure to RRT is risk factor for major gastrointestinal (GI) bleeding [7–10], clarifying whether clinically important upper GI bleeding can be prevented with proton pump inhibitors (PPIs) during critical illness is clinically relevant. Two recent international randomized trials [11, 12] and an updated meta-analysis [13] have supported the use of PPIs in the ICU to reduce the risk of clinically important upper GI bleeding in critically ill patients. However, PPIs have been reportedly associated with AKI and other adverse kidney outcomes in some populations [14–20], although not clearly in critically ill patients. PPIs are also commonly prescribed for outpatients with kidney failure [21–23].

One of the two aforementioned international randomized trials was the Re-evaluating the Inhibition of Stress Erosions (REVISE) multicenter randomized trial, which found pantoprazole to significantly lower the risk of clinically important upper GI bleeding, with no effect on mortality [11]. Nevertheless, concerns have emerged about PPI exposure during critical illness, due to a possible heightened risk of mortality among the most seriously ill patients based on subgroup analyses of two prior trials [12, 24]. Further, it is not clear whether and how kidney failure predisposes a patient to bleeding – including whether it is due to platelet dysfunction or anticoagulation associated with RRT [7] and whether PPIs are more or less effective in this population. A previous substudy [25] of the Stress Ulcer Prophylaxis in the ICU (SUPICU) trial [12] found no statistically significant interactions regarding the effect of pantoprazole between patients with versus without RRT at baseline, although maintenance RRT was not evaluated separately. Whether RRT before critical illness has any modifying effect on the effectiveness of pantoprazole at preventing clinically important upper GI bleeding in the ICU merits further study. To address this question, we have planned a *post hoc* secondary analysis of an international stress ulcer prophylaxis trial.

## METHODOLOGY

### Study Design & Objectives

This study is a *post hoc* analysis of the REVISE Trial (ClinicalTrials.gov: NCT03374800) [11].

The primary objective of this study is to evaluate whether any RRT at baseline (e.g., maintenance RRT in the outpatient setting, or RRT initiated in the hospital [intermittent hemodialysis, continuous renal replacement therapy, sustained low efficiency dialysis, or peritoneal dialysis]) modifies the treatment effect of pantoprazole on clinically important upper GI bleeding and 90-day mortality.

The secondary objective is to evaluate specifically whether maintenance RRT (intermittent hemodialysis or peritoneal dialysis) [26] in an outpatient setting before the index hospital admission modifies the effect of pantoprazole on clinically important upper GI bleeding and 90-day mortality. Secondary outcomes for both the primary and secondary objectives are patient important upper GI bleeding, and mortality in the ICU and in the hospital.

### Summary of the REVISE Trial

REVISE was a blinded randomized controlled trial of 4,821 invasively ventilated patients that evaluated the efficacy and safety of pantoprazole for stress ulcer prophylaxis. Patients aged ≥18 years of age admitted to the ICU and expected to remain invasively ventilated beyond the day after enrollment were eligible. Exclusion criteria were invasive ventilation for >72 hours at the time of screening, receipt of >1 day of PPI or histamine-2-receptor-antagonist therapy in the ICU, dual antiplatelet therapy or combined antiplatelet and therapeutic anticoagulation, active upper GI bleeding, and contraindications to PPI. Patients were randomly assigned to receive intravenous pantoprazole 40 mg once daily or placebo while invasively ventilated in the ICU for up to 90 days. Study drug was discontinued if contraindications or clinical indications to the use of PPIs arose. The protocol [27], statistical analysis plan [28], and approach during the pandemic [29] were reported previously. We will use baseline and daily data collected in the REVISE trial from all 68 centers in Australia, Brazil, Canada, Kuwait, Pakistan, Saudi Arabia, United Kingdom, and the United States. No additional data will be required for this analysis.

### Outcomes

The primary efficacy outcome for this study is the same as the main REVISE trial - clinically important upper GI bleeding in the ICU, identified locally as overt GI bleeding with evidence of hemodynamic compromise or leading to therapeutic interventions in the ICU (or resulted in readmission to the ICU during the index hospital stay) up to 90 days after randomization [11, 27]. Similarly, the primary safety outcome for this study is death from any cause at 90 days [11, 27]. Secondary outcomes of interest for both the primary and secondary objective will include patient important upper GI bleeding in the ICU, ICU mortality, and hospital mortality. The definition of patient important upper GI bleeding was previously established through a mixed methods study of ICU survivors and family members [30]. Upper GI bleeding was considered patient-important if it required a single blood transfusion, vasopressor treatment, diagnostic endoscopy, CT-angiography, surgery, or if it resulted in death, disability, or prolonged hospitalization [31]. These criteria were applied to the REVISE database up to 90 days after randomization.

### Hypotheses

Our *a priori* hypothesis is that use of PPI, as compared with placebo, will reduce the occurrence of clinically important upper GI bleeding among patients receiving any RRT at baseline, but will have no effect on 90-day mortality. We further hypothesize an effect modification – that is, that the effect of PPIs on reducing upper GI bleeding events will be greater than the treatment effect observed across the remaining cohort, even in the context of a higher prevalence of pre-hospital PPI use among patients receiving maintenance RRT than other patients.

Regarding the secondary objective, we hypothesize no subgroup effects.

### Statistical Analysis Plan

We will use descriptive statistics including measures of central tendency and dispersion as appropriate, and counts and percentages. We also analyze the duration of RRT amongst those newly receiving RRT in the ICU in days.

To address the primary objective, we will conduct a Cox proportional hazards time-to-event analysis for the primary and secondary outcomes. Each model will include the randomized exposure of pantoprazole vs placebo, receipt of pre-hospital acid suppression (as in the original trial analyses), and RRT (either maintenance RRT in the outpatient setting [n=90], or RRT initiated in the hospital (intermittent hemodialysis, continuous renal replacement therapy, sustained low efficiency dialysis, or peritoneal dialysis) [n=255]), and its interaction with randomized exposure. Additionally, mortality outcomes will be adjusted for baseline illness severity as measured by the Acute Physiology and Chronic Health Evaluation (APACHE) II score [32].

Model assumptions and goodness-of-fit testing, including the proportional hazards assumption, will be performed graphically. Results will be reported as hazard ratios (HRs), with associated 95% confidence intervals (CIs) around the effect of pantoprazole on each outcome relative to placebo for each sub-group). Interaction p-values will be reported. If evidence of a sub-group interaction is present (p-value for interaction <0.1), we will assess subgroup credibility using the ICEMAN instrument [33].

To address the secondary objective, we will perform the same Cox proportional hazards time-to-event analysis on the same outcomes; however, the definition of RRT will be maintenance RRT (intermittent hemodialysis or peritoneal dialysis) in an outpatient setting before the index hospital admission; n=90]). Similarly, we will report the analyses as HRs with 95% confidence intervals and an interaction p-value. All P-values will be two-sided, with P<0.05 considered statistically significant.

Patients will be evaluated in the group to which they had been randomly allocated.

### Ethics & Safety Considerations

Participant informed consent processes, trial registration (NCT03374800), and trial oversight have been previously reported [27–29]. Patients and/or their substitute decision-makers gave informed consent for participation in the REVISE trial. Consent procedures included *a priori* informed consent, or deferred consent (e.g., consent to continue); an opt-out model was used in one center. The trial protocol was approved by ethics review boards and relevant regulatory agencies at each site. There was no additional ethics review for this secondary analysis of previously collected, de-identified data.

### Dissemination

We plan to publish the results of this study in a peer-reviewed journal, The study team, alongside collaborative team members from the original REVISE trial, will lead local, national, and international knowledge translation efforts in the form of presentation at conferences, local grand rounds, and other academic settings. We will also create lay language summaries, infographics and briefs for traditional and social media platforms.

## DISCUSSION

This study is a subgroup analysis using a recently published international randomized trial database of stress ulcer prophylaxis. We will evaluate whether there is a modification of the effect of pantoprazole according to baseline RRT exposure considering the outcomes of clinically important upper GI bleeding, patient important GI bleeding, 90-day, ICU and hospital mortality.

This study has several strengths, including the large number of international participants and ascertainment of RRT status pre-hospital and within hospital. This focus on critically ill patients receiving two modes of advanced life support - invasive mechanical ventilation and RRT - will explore the effect of a commonly administered pharmacological intervention to prevent upper GI bleeding. The analysis is pre-planned as outlined herein.

Limitations of this study include the *post hoc* nature of the subgroup analysis, which reduces the credibility of any finding of interaction. Confidence intervals may be wide due around some estimates to small sample sizes. Some RRT exposure will be too uncommon to allow detailed analysis of mode (e.g., peritoneal dialysis). While we examine the specific effects of renal insufficiency requiring RRT, we acknowledge that renal insufficiency exists on a spectrum. We did not collect the dose of RRT-associated anticoagulation or citrate. Also, when adjusting for acid suppression pre-hospital, we will not be able to distinguish between either a PPI or histamine-2-receptor antagonist prescription as these drugs were distinguished in hospital but not pre-hospital.

## CONCLUSION

This study will explore whether pantoprazole is more, less, or similarly effective among critically ill patients with kidney failure severe enough to receive RRT in the hospital before ICU admission or as an outpatient only, relative to other patients. The resulting evidence will have direct implications for critical care practice, particularly for patients exposed to RRT at risk of stress-related GI ulceration in the ICU.

## Data Availability

As a protocol for a proposed study, there is no data referred to in the manuscript.

## ACKNOWLEGEMENTS

We appreciate the patients and family members who participated in the REVISE trial. We are grateful for the collaboration of all the Research Coordinators and Site Investigators. REVISE was designed by the REVISE Steering Committee including National and International Management Committees, the REVISE Investigators and Research Coordinators, the Canadian Critical Care Trials Group, and the Australian and New Zealand Intensive Care Society Clinical Trials Group. We are grateful to the Methods Center teams at McMaster University (Lisa Buckingham, France Clarke, Mary Copland, Karlo Matic, Ashley Sawyer, Alyson Takaoka) and The George Institute (Fatima Butt, Anna Cheng, Conrad Nangla, Fiona Osborne, Tina Schneider, Isabella Schoeler, Anna Tippet) for their expertise. Sean Bagshaw is supported by a Canada Research Chair in Critical Care Outcomes and Systems Evaluation. Chris Yarnell is supported by the Northpine Professorship in Critical Care Research, Scarborough Health Network Foundation; Canadian Critical Care Trials Group Early Researcher Award; Interdepartmental Division of Critical Care Medicine Scholar Award; and the National Sanitarium Association Scholar Award. Deborah Cook is supported by the St. Joseph’s Healthcare/McMaster University Critical Care Chair.

## AUTHOR CONTRIBUTIONS - to be updated

### Concept and Design

A Thabane, O Rewa, D Heels-Ansdell, SM Bagshaw, R Jeong, M Borthwick, NKJ Adhikari, FG Zampieri, A Kuriyama, N Zytaruk, D Cook

### Acquisition, Analysis, or Interpretation of Data

A Thabane, D Heels-Ansdell

### Drafting of the Manuscript

A Thabane, D Cook, D Heels-Ansdell, SM Bagshaw, R Jeong, M Borthwick, NKJ Adhikari, A Kuriyama, J Marshall

### Critical Revision of the Manuscript for Important Intellectual Content

All coauthors

### Obtained Funding for this Secondary Analysis of the REVISE Grant

Not applicable

### Administrative, Technical, or Material Support

D Heels-Ansdell, A Thabane, N Zytaruk

### Data Integrity

D Heels-Ansdell, N Zytaruk, D Cook

